# Genome-Wide Association Studies and Deep-Learning Functional Annotation of Opioid Use Disorder across Three Ancestries in the *All of Us* Research Program

**DOI:** 10.64898/2026.07.15.26358096

**Authors:** Shaopeng Gu, Dan Petrovitch, O. Trent Hall, Joshua W. Lambert, Rachel L. Kember, Noor A. Nahid, Qin Ma, Jon E. Sprague, Caitrin W. McDonough, Julie A. Johnson

**Affiliations:** Department of Biomedical Informatics, College of Medicine, The Ohio State University, Columbus, OH, USA; Pelotonia Institute for Immuno-Oncology, James Comprehensive Cancer Center, The Ohio State University, Columbus, OH, USA; Department of Psychological Sciences, Texas Tech University, Lubbock, TX, USA; Faillace Department of Psychiatry and Behavioral Sciences, University of Texas Health Science Center at Houston, Houston, TX, USA; Department of Psychiatry and Behavioral Health, College of Medicine, The Ohio State University, Columbus, OH, USA; College of Nursing, University of Cincinnati, Cincinnati, OH, USA; Department of Psychiatry, University of Pennsylvania Perelman School of Medicine, Philadelphia, PA, USA; The Ohio Attorney General’s Center for the Future of Forensic Science, Bowling Green State University, Bowling Green, OH, USA; Department of Pharmacotherapy and Translational Research, College of Pharmacy, University of Florida, Gainesville, FL, USA; Clinical and Translational Science Institute, The Ohio State University, Columbus, OH, USA; Department of Internal Medicine and Department of Pharmaceutics & Pharmacology, College of Medicine, The Ohio State University, Columbus, OH, USA

**Keywords:** opioid use disorder, genome-wide association study, genetic ancestry, All of Us Research Program, AlphaGenome, functional genomics

## Abstract

**Background:** Opioid use disorder (OUD) is heritable, yet most genome-wide association studies (GWAS) have focused on European populations, leaving the genetic architecture of OUD in non- European populations underexplored.

**Methods:** We conducted GWAS of OUD across three ancestries using electronic health records and genomic data from 52,357 *All of Us* Research Program participants (8,912 cases; 43,445 matched opioid-exposed controls; 48.5% female). Participants were stratified into European (EUR), African (AFR), and Admixed American (AMR) ancestry groups for logistic regression GWAS, with independent replication in the Million Veteran Program. We then applied the deep- learning model AlphaGenome to predict the tissue-specific transcriptomic and splicing consequences of top risk variants across 13 reward-pathway brain regions.

**Results:** We identified and replicated a novel *DDX6* risk locus, alongside established *OPRM1* and *FURIN* signals. AlphaGenome predicted the *DDX6* regulatory allele downregulates the stress-resistance gene *FOXR1* in the nucleus accumbens, while the protective *OPRM1* variant (rs1799971) upregulates *OPRM1* expression across reward networks. Other signals of interest included *IL6R* and *SHISA9* (EUR); *GHR* (AFR); and *ASTN2* (AMR).

**Conclusions:** This study identifies *DDX6* as a novel OUD risk locus, replicates associations with *OPRM1* and *FURIN*, and highlights biologically plausible ancestry-specific signals in AFR and AMR populations. We also replicated top variants in an independent population. Finally, integrating GWAS with deep-learning annotations provides specific, localized biological hypotheses to guide future experimental validation and targeted therapeutics.

## Introduction

The opioid crisis is a critical public health challenge. In 2024, more than 4.8 million people experienced opioid use disorder (OUD) within the past year in the U.S. (1), contributing to over 54,000 opioid-involved overdose deaths (2). This lethal, costly burden has persisted despite significant preventive efforts, the development of pharmacological and adjunctive behavioral interventions for OUD, and advances in our understanding of the neurobiology of OUD (3).

Addressing this burden will require continued progress in prevention and treatment as well as a more complete picture of the biological, behavioral, and psychosocial etiologies of OUD. The genome-wide association study (GWAS) is a powerful method for advancing these aims. By testing associations between OUD and single nucleotide polymorphisms (SNPs) and indels across the entire genome, GWAS may help identify clinically relevant genetic markers for OUD risk, suggest novel targets for drug development, improve our knowledge of the biology of OUD, and inform models designed to make causal inferences about non-biological risk factors for OUD (4).

Despite estimates from twin and family studies that approximately 50% of OUD risk is heritable (5–7), OUD GWAS have struggled to identify consistent risk loci. Prior studies have implicated a wide variety of plausible genes, including *RGMA* (8), *KCNC1*, *KCNG2* (9), *CNIH3* (10), *PPP6C* (11), and *KDM4A* (12), among others. Attempts to replicate *RGMA*, *KCNG2*, and *CNIH3* using independent datasets have been unsuccessful (13,14).

*OPRM1* and *FURIN* are examples of genes that have been replicated in OUD GWAS. As the gene encoding the μ-opioid receptor (the main protein target of opioids) (3), *OPRM1* has long been a target of candidate-gene studies focused on OUD and related phenotypes (15–17). Zhou et al. (18) reported the first genome-wide significant *OPRM1* signal at rs1799971 (A118G) from meta-analyses of European American U.S. Veterans from the Million Veterans Program (MVP) and various non-Veteran samples, with the minor allele (G) showing a small protective effect (β = −0.066, *p* = 1.51 × 10^-8^). Kember et al. (19) replicated this finding after extending their analysis of the MVP to include Hispanic American Veterans and loosening their definition of cases. Deak et al. (20) also replicated this finding by meta-analyzing these data (including all cases from Zhou et al.’s study) with several additional institutional datasets. Beyond rs1799971, they identified a cross-ancestry signal at *OPRM1* rs9478500. Gaddis et al. (11) similarly found genome-wide significance for rs9478500 using genomic structural equation modeling to combine novel data with recycled summary statistics from the MVP and other datasets, while their haplotype analyses suggested that rs1799971 may not be the causal variant. Notably, rs1799971 has not reached genome-wide significance in any of the AFR-only meta-analyses to date.

Several of these studies have also converged on *FURIN*, which encodes furin, an endoprotease expressed in many tissues (including the brain) that catalyzes activation of numerous proprotein substrates (21–23). For instance, both SNP- and gene-based approaches have identified associations between OUD and *FURIN* variants rs17514846 (11,20), rs11372849 (11,20), rs4702, and rs3759929 (19,24). However, although these studies are often cited to provide convergent evidence for the role of *OPRM1* and *FURIN* in OUD, they do not represent fully independent replications due to the substantial reuse of the same data across meta-analyses.

Beyond the inconsistency of findings (outside of the literature’s overreliance on the same datasets), many extant OUD GWAS have limited generalizability for non-EUR individuals. Most loci have been identified among participants of European ancestry (EUR), while sample sizes for participants of African ancestry (AFR) have been notably smaller. There is an even starker gap in the literature for individuals of Admixed American ancestry (AMR) (a group that includes many, but not exclusively, Hispanic individuals) (24). To date, only Kember et al. (19) have published an OUD GWAS including a Hispanic American sample, finding a Hispanic-specific genome-wide significant risk signal near *MRS2*. As such, Deak and colleagues explicitly called for “purposeful recruitment of AFR and other non-EUR OUD subjects” for OUD GWAS (20).

Compounding the issue of limited ancestral representation in OUD GWAS, the MVP is predominantly composed of males, reflecting the historically high proportion of males in the U.S. military (25). Because the MVP is such a large sample with so many OUD cases, it often disproportionately contributes to OUD GWAS meta-analyses (18,19). As a result, there is a high proportion of males contributing to the evidence base for genetic risk factors for OUD.

The NIH *All of Us* Research Program is a large, ancestrally diverse convenience sample of U.S. adults (26,27). This makes the *All of Us* data remarkably well positioned to address the limited ancestral representation and lack of truly independent cohorts characterizing extant OUD GWAS. Since May of 2018, participants have enrolled via a national, multimodal recruitment network of clinical and community-based partners (28,29). Participants are asked to provide permission to link their electronic health records (EHRs) with their genomic and self-report survey data (30). Crucially, data from *All of Us* have not yet contributed to any published OUD GWAS meta-analyses, although investigators have used the dataset to investigate the genomics of several related outcomes (e.g., opioid use, other substance use disorders) (31–33).

Therefore, we performed a GWAS to explore genetic variants associated with OUD among EUR, AFR, and AMR participants from the *All of Us* Research Program. We successfully replicated several of our genome-wide significant SNPs using summary statistics from a large, independent, multi-ancestry OUD GWAS (19). Finally, we used the deep-learning model AlphaGenome to evaluate the potential functional impacts of these variants (34). To our knowledge, our study is the first OUD GWAS conducted in the *All of Us* dataset, the first OUD GWAS to use AlphaGenome for functional annotation and variant interpretation, and only the second OUD GWAS conducted in an AMR population (19). Our findings revealed novel genetic associations for each ancestry group, pointing toward biological pathways that may contribute to OUD.

## Methods and Materials

We used large language models (LLMs) throughout the research process and disclose that use here in accordance with the journal’s policy on AI technologies (35), including various models provided by Anthropic (San Francisco, CA), OpenAI (San Francisco, CA), and Google (Alphabet, Inc., Mountain View, CA). These tools were used to assist with data analysis (e.g., drafting analytic code), writing (e.g., editing, proofreading, revising, formatting), and literature review (e.g., searching for relevant articles to read). We reviewed and revised model outputs for accuracy prior to use and take responsibility for the content of this article. We complied with the *All of Us* Research Program’s policies, and no participant-level data were uploaded to LLMs. Additional details are available upon request.

### Setting and Participants

This study was a secondary analysis of de-identified data (registered and controlled tiers) from the All of Us Research Program. We accessed select EHR, short-read Whole Genome Sequencing (srWGS), and survey data from *All of Us* Version 8, which reports a cutoff date of October 1, 2023 (36). Based on previous literature (18,19), OUD cases were participants with available srWGS data and relevant OUD ICD-9/-10 codes in their EHR from ≥ 1 inpatient or ≥ 2 outpatient visits. Table S1 lists the diagnostic codes used to extract standardized clinical concepts for opioid abuse or dependence in *All of Us* (37,38). We restricted controls to participants with available srWGS data and history of opioid exposure documented in their EHR. Specifically, we defined controls as participants with record of exposure to any drug from a list of 17 prescription opioids with ≥ 1 inpatient or ≥ 2 outpatient visits, excluding those with any evidence of OUD, OUD pharmacotherapies (buprenorphine, methadone, naltrexone), or cancer-related diagnoses. This yielded 8,987 cases and 122,699 controls prior to stratification by ancestry (see Figure 1).

**Figure 1.**
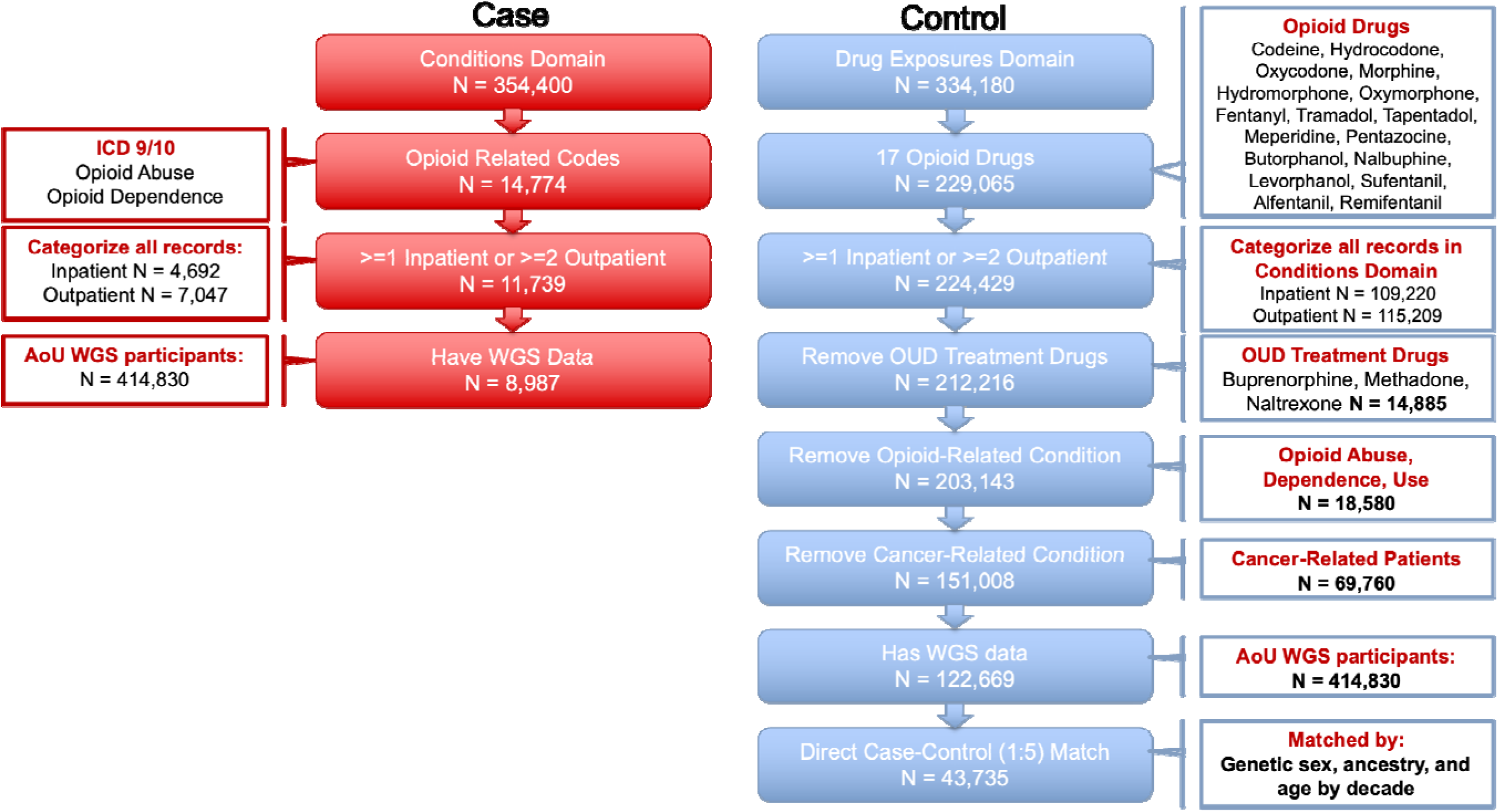
Flowchart of data pre-processing and EHR-based case and control definitions for opioid use disorder prior to ancestry grouping. *Note. N*’s within red and blue boxes (under Case and Control) reflect sample sizes remaining at each stage of the pre-processing pipeline. *N*’s within white boxes (on the left and right sides of the figure) reflect sample sizes for each characteristic across the entire All of Us Version 8 dataset (except for the two boxes on the categorization of records by inpatient/outpatient codes). EHR = electronic health record, AoU = All of Us, WGS = whole-genome sequencing

We then partitioned these cases and controls into three ancestry groups based on the categorical indicators of genetic ancestry provided by *All of Us*: EUR, AFR, or AMR^1^ (39). For each ancestry group, we performed case-control matching up to a 1:5 case-control ratio (while retaining all cases) based on genetic sex and age deciles. This yielded final, ancestry-specific analytic cohorts of 27,834 EUR (*n* = 4,725 cases), 15,103 AFR (*n* = 2,617 cases), and 9,420 AMR participants (*n* = 1,570 cases). These translate to effective sample sizes of *N* = 15,692 EUR, *N* = 8,654 AFR, and *N* = 5,233 AMR participants, using a recommended heuristic (18,40).

### Quality Control and Discovery GWAS

In the *All of Us* Research Program, blood- or saliva-derived DNA samples are sequenced by one of several Genome Centers that use harmonized wet lab protocols and quality control methodologies. Comprehensive details on sample handling, sequencing, and quality control steps are available elsewhere (30,39,41). We utilized the Allele Count/Allele Frequency threshold callset (multiallelic split dataset), which is an *All of Us*-provided, reduced set of more than 116 million common variants (covering over 56 million loci) with either a minor allele frequency above 1% or a minor allele count above 100 in any ancestry subgroup (42). We applied additional quality control steps within each ancestry group by excluding variants deviating from Hardy-Weinberg equilibrium (threshold: *p* < 1 x 10^-6^) and those with minor allele frequencies below 1%. We then performed GWAS within each ancestry subgroup, implementing unconditional binary logistic regression models via the Python package *Hail*, version 0.2 (43), within the *All of Us* Researcher Workbench. For each model, covariates were initially the first 10 *All of Us*-provided ancestry principal components (PCs), genetic sex, and age, as appropriate for matched designs (44). We set *p* < 5 x 10^-8^ as the genome-wide significance threshold (α), per conventions for GWAS (45).

#### Principal Components Analysis

QQ plots of the resulting GWAS data revealed high levels of genomic inflation in the AMR (λ_GC_ = 1.29) and EUR (λ_GC_= 1.12) ancestry groups. We thus performed our own principal components analysis (PCA) within each ancestry group to capture the more fine-grained population substructure in the EUR and AMR ancestry groups. Specifically, we selected high- quality, common variants based on the following criteria: minor allele frequency > 0.1, Hardy- Weinberg equilibrium *p* > 0.001, and variant call rate > 99.5%. From the pool of variants meeting those criteria, we randomly sampled approximately 150,000 variants then pruned those in linkage disequilibrium (based on: *R^2^*< 0.1, window size = 100 kb). We ran a PCA on this finalized set of relatively independent signals using *Hail*. We then reran our GWAS using the first 10 PCs from our ancestry-specific PCA as covariates for each model, rather than *All of Us*- provided PCs, along with genetic sex and age.

The selection of covariates and reported models differed by ancestry group. For AMR (initial λ_GC_ = 1.29, attenuated to λ_GC_ = 1.04 with our PCs), we only present models that used our calculated PCs. For EUR (initial λ_GC_ = 1.12, attenuated to λ_GC_ = 1.07 with our PCs), we present both sets of results: models using *All of Us*-provided PCs in the primary manuscript and those using our calculated PCs in the Supplementary Information. For AFR, the inflation was already minimal using the *All of Us*-provided PCs (λ_GC_= 1.01), so we only present one model without any AFR-specific PC calculation.

### Replication in Independent Meta-Analysis

We sought to replicate our findings with available summary statistics in the independent cross-ancestry meta-analysis conducted by Kember et al. (2022) (19). Each variant of interest was queried in the specific ancestral population of our signal. The discovery data reported herein were derived from genome sequencing, whereas the replication sample had SNP array data, so many of our top signals were not available in the summary statistics from the Kember et al. paper. 18 SNPs in EUR (representing 2 signals), 1 SNP in AFR, and 11 SNPs (representing 2 signals) in AMR were tested for replication. Thus, we tested 5 independent signals, so significant replication was defined as P < 0.01 and a directionally consistent finding.

We also performed cross-ancestry look-ups of our top signals in the other two ancestry groups. This resulted in 2 signals from *DCUN1D4* and *ASTN2* evaluated in EUR, 6 signals from *IL6R, CYP3A54P, FKBP4P7, DDX6, RNU6-1230P,* and *ASTN2* in AFR, and 6 signals from *IL6R, CYP3A54P, FKBP4P7, DDX6, SHISA9,* and AF274853.1 in AMR (Table 2).

We also attempted to replicate select SNPs from previous OUD GWAS literature using our summary statistics, including OPRM1 rs1799971, rs9478500 (11,18), and FURIN rs3759929, rs17514846, rs11372849, rs4702 (11,19,20), with a significance threshold P < 0.0083 (0.05/6) and a directionally consistent finding.

### Functional Annotation and Variant Interpretation

Finally, to elucidate the potential functional consequences of variants identified in our GWAS, we used AlphaGenome, a recently released, multimodal sequence-to-function deep-learning model that balances the ability to make high-resolution predictions with the ability to accept longer input sequences (up to 1,048,576 base pairs) (34). We applied this model to make predictions about the potential functional consequences of our SNP-level findings on different modalities, such as gene expression, and splicing in brain tissues relevant to OUD and the brain disease model of addiction. Specific tissues (and their Uberon identifiers) were the nucleus accumbens (NAc; 0001882), including the NAc shell (0012171) and core (0012170), dorsolateral prefrontal cortex (dlPFC; 0009834), striatum (0002435), ventral tegmental area (VTA; 0002143), amygdala (0001876), hippocampus (cornu ammonis; 0001954), orbitofrontal cortex (OFC: 0002267), anterior cingulate cortex (0009835), insula (0002022), bed nucleus of the stria terminalis (BNST; 0001880), and habenula (0001904). Notably, while gene expression tracking accounts for these distinct anatomical regions, AlphaGenome’s core splice site prediction operates as a sequence-based, tissue-agnostic modality, thereby producing a unified prediction across all specified Uberon identifiers.

## Results

### Demographics and Matching

The final study cohort consisted of 52,357 participants, comprising 8,912 OUD cases and 43,445 opioid-exposed controls. See Table 1 for basic sociodemographic information on cases and controls, stratified by ancestry. Tables S2 and S3 provide descriptive statistics on additional self-reported demographic, health, and substance use variables, derived from the basic, overall health, health care access and utilization, and lifestyle surveys available from *All of Us* (46). Beyond our goal of providing a more comprehensive picture of our analytic samples, these data can provide some evidence of the construct validity of our EHR-based OUD operationalization by demonstrating that cases generally exhibited expected associations with known correlates of OUD (47,48).

**Table 1.** Sample sizes and basic demographics for cases and controls by genetically informed ancestry group.

| Variable | EUR |  | AFR |  | AMR |  |
| --- | --- | --- | --- | --- | --- | --- |
|  | Case | Control | Case | Control | Case | Control |
| *n* | 4,725 | 23,109 | 2,617 | 12,486 | 1,570 | 7,850 |
| Age (mean ± *SD*) | 53.28 ± 14.0 | 53.72 ± 14.2 | 59.09 ± 11.7 | 55.05 ± 14.8 | 49.83 ± 12.9 | 49.83 ± 13.3 |
| Genetic sex: female | 2,454 (51.9%) | 12,270 (53.1%) | 1,119 (42.8%) | 5,595 (44.8%) | 663 (42.2%) | 3,315 (42.2%) |
| Genetic sex: male | 2,271 (48.1%) | 10,839 (46.9%) | 1,498 (57.2%) | 6,891 (55.2%) | 907 (57.8%) | 4,535 (57.8%) |
| Ethnicity<sup>a</sup> |  |  |  |  |  |  |
| Hispanic/Latino | 60 (1.3%) | 337 (1.5%) | 112 (4.3%) | 705 (5.6%) | 1,270 (80.9%) | 6,799 (86.6%) |
| Not Hispanic/Latino | 4,468 (94.6%) | 22,288 (96.4%) | 2,375 (90.8%) | 11,324 (90.7%) | 241 (15.4%) | 877 (11.2%) |

*Note.* OUD = opioid use disorder, EUR = European ancestry, AFR = African ancestry, AMR = Admixed American ancestry, Case = OUD-positive, Control = OUD-negative, *SD* = standard deviation. Statistics are count (percentage) unless otherwise indicated. <sup>a</sup>Some participants selected *Skip* or *Prefer Not To Answer*.

### Discovery of Ancestry-Specific OUD Risk Loci

Our three GWAS identified a total of 60 genome-wide significant variants (*p* < 5 x 10^-8^) distributed across 12 independent loci, Table 2. Top signals were not consistent across the ancestry groups. All Manhattan and Q-Q plots are included in the Supplementary document.

**Table 2.** Summary of the 12 genome-wide significant independent signals of opioid use disorder by ancestry and 3 well-known signals evaluated for replication.

| SNP | Gene | Ancestry Group |  |  |  |  |  |  |  |  |
| --- | --- | --- | --- | --- | --- | --- | --- | --- | --- | --- |
|  |  | EUR |  |  | AFR |  |  | AMR |  |  |
|  |  | *p* | *OR* | *ALT*<br>*AF* | *p* | *OR* | *ALT*<br>*AF* | *p* | *OR* | *ALT*<br>*AF* |
| rs7411976<br>1:154409967:A>C | IL6R | **6.8E-09** | **1.64** | **0.014** | 5.2E-01 | 1.05 | 0.052 | 8.9E-02 | 0.76 | 0.020 |
| rs1323472410<br>7:4407902:AT>A | CYP3A54P | **3.5E-08** | **1.49** | **0.527** | 2.8E-01 | 1.12 | 0.561 | 9.5E-01 | 0.99 | 0.572 |
| rs1164905391<br>9:41624733:T>A | AL591926.7 | **1.6E-11** | **0.66** | **0.068** | - | - | - | - | - | - |
| rs1344710397<br>9:62709092:A>G | FKBP4P7 | **4.4E-08** | **1.55** | **0.283** | 5.9E-01 | 1.06 | 0.265 | 9.6E-02 | 1.10 | 0.294 |
| rs472419<br>11:118722509:C>T | DDX6 | **1.0E-08** | **0.87** | **0.621** | 8.1E-01 | 1.00 | 0.441 | 7.1E-01 | 1.01 | 0.717 |
| rs1202910945<br>16:12970492:TATATA>T | SHISA9 | **7.0E-09** | **1.46** | **0.262** | - | - | - | 4.0E-01 | 1.12 | 0.209 |
| rs5924907<br>X:151344720:G>C | AF274853.1 | **7.0E-09** | **1.51** | **0.034** | - | - | - | 2.6E-01 | 1.17 | 0.026 |
| rs116570345<br>5:42658399:G>T | GHR | - | - | - | **3.6E-08** | **2.01** | **0.012** | - | - | - |
| rs376702299<br>14:91309609:CA>C | CCDC88C | - | - | - | **3.9E-08** | **1.84** | **0.018** | - | - | - |
| rs17473535<br>4:51888241:A>G | DCUN1D4 | 7.1E-01 | 0.97 | 0.032 | - | - | - | **3.4E-08** | **2.24** | **0.015** |
| rs6831654<br>4:149837132:A>G | RNU6-1230P | - | - | - | 5.2E-02 | 0.90 | 0.125 | **4.5E-08** | **2.07** | **0.018** |
| rs6478237<br>9:116430237:A>C | ASTN2 | 1.7E-01 | 1.04 | 0.773 | 8.4E-01 | 0.99 | 0.818 | **5.8E-09** | **1.41** | **0.817** |
| Replication of previously documented OUD variants |  |  |  |  |  |  |  |  |  |  |
| rs1799971<br>6:154039662:A>G | OPRM1 | **9.0E-06** | **0.85** | **0.128** | 2.0E-01 | 0.88 | 0.030 | **2.3E-03** | **0.83** | **0.176** |
| rs9478500<br>6:154057088:T>C | OPRM1 | **3.5E-04** | **1.11** | **0.174** | 3.6E-01 | 1.03 | 0.183 | 6.4E-02 | 1.12 | 0.126 |
| rs3759929<br>15:90866779:G>T | FURIN | **9.7E-03** | **0.94** | **0.594** | **6.2E-03** | **0.90** | **0.754** | - | - | - |

*Note.* Probability (*p*) and odds ratio (*OR*) are shown for each independent signal in the European ancestry (EUR), African ancestry (AFR), and Admixed American ancestry (AMR) groups. SNP = single-nucleotide polymorphism; ALT AF = alternate allele frequency; QC = quality control; MAF = minor allele frequency; HWE = Hardy-Weinberg equilibrium. A dash indicates that the SNP was filtered by QC in that ancestry (MAF < 1% or HWE $p < 1 \times 10^{-6}$ ). Signals meeting the defined significance thresholds are shown in bold.

### European Ancestry (EUR) GWAS

In the EUR cohort, 39 variants reached genome-wide significance, representing seven independent signals, of which three had multiple significant SNPs/indels in the region. These included *IL6R* (Interleukin-6 Receptor) rs7411976 (*p =* 6.79 x 10^-9^)*. DDX6* had a dense cluster of 25 significant variants. Most notably, this signal includes the missense variant rs472419 (*p =* 1.01 x 10^-8^).; which alters the amino acid sequence of the protein (Ala314Pro/Thr). Finally, multiple signals were noted in *SHISA9* with the most significant being rs1202910945 (*p =* 7.04 x 10^-9^).

#### Replication of Known OUD Risk Loci of EUR

Consistent with findings from the Million Veteran Program (MVP) (18,19), we replicated the association of the *OPRM1* variant rs1799971 (*p =* 9.0 x 10^-6^,OR =0.85), rs9478500 (*p =* 3.5 x 10^-4^,OR = 1.11), and the *FURIN* variant rs3759929 (*p =* 9.7 x 10^-3^,OR = 0.94) in the EUR cohort, Table 2.

### African Ancestry (AFR) GWAS

Despite being a smaller sample size compared to the EUR group, the AFR cohort yielded two genome-wide significant signals, which included signals in *GHR*(*P* = 3.62 x 10^-8^;rs116570345) and *CCDC88C* (*P* = 3.98 x 10^-8^;rs376702299). We replicated the association of the *FURIN* variant rs3759929 (*P* = 6.2 x 10^-3^,*OR*=0.9) in the AFR cohort, Table 2.

### Admixed American Ancestry (AMR) GWAS

In the AMR cohort, 19 variants reached genome-wide significance, coalescing into 3 independent signals. *ASTN2* had 15 significant variants with the top SNP being rs6478237 (*p =* 5.28 x 10^-9^)*. DCUN1D4* rs17473535 (*p =* 3.42 x 10^-8^). and *RNU6-1230P* rs6831654 (*p =* 4.50 x 10^-8^). were also identified. We replicated the association of the *OPRM1* variant rs1799971 (*P* = 2.3 x 10^-3^,*OR*=0.83) in the AMR cohort, Table 2.

### External Replication of OUD Loci

Based on available SNP-level data in the replication cohort, we were able to attempt replication of the signals in *IL6R* and *DDX6* in EUR, *GHR* in AFR, and *DCUN1D4* and *ASTN2* in AMR. All twelve variants in *DDX6* passed the replication criteria (*P*<0.001), with the functional missense variant rs472419 (Ala314Pro/Thr) in *DDX6* having a *P* = 0.0042 (*OR* 1.03). *IL6R* in EUR did not replicate, nor did any signals in AFR and AMR.

### Functional Annotation of Ancestry-Specific Loci using AlphaGenome

We used the AlphaGenome sequence-to-function deep-learning model to predict the transcriptomic and regulatory consequences of the fine-mapped variants.

In the EUR cohort, the *DDX6* variant (chr11:118713914:AT>A) is predicted to downregulate *FOXR1* expression within the nucleus accumbens basal ganglia (log-fold change = -0.166; quantile = -0.9999) (Figure 2A). The *IL6R* variant (chr1:154411947:CT>C) is predicted to impact alternative splicing, affecting specific splice junctions (raw score = 1.127; quantile = 0.9997), overall splice site usage (raw score = 0.250; quantile = 0.9996), and donor splice site probability (raw score = 0.131; quantile = 0.992) (Figure 2B). The protective *OPRM1* variant rs1799971 (chr6:154039662:A>G) is predicted to upregulate *OPRM1* expression in the amygdala, nucleus accumbens, and anterior cingulate cortex (log-fold change = 0.034; quantile = 0.9996), as well as in the dorsolateral prefrontal cortex and hippocampus (Figure 2C).

**Figure 2.**
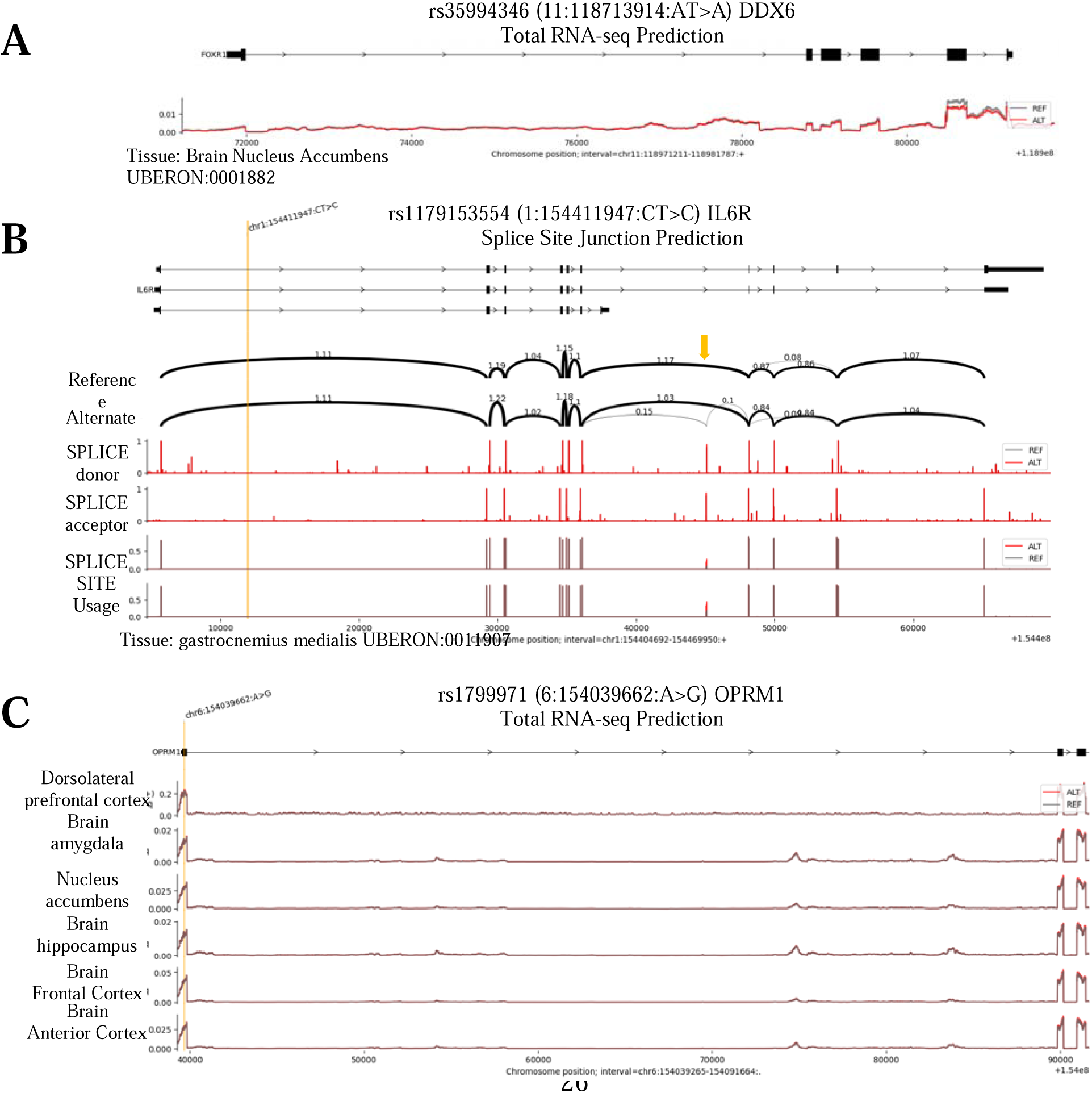
In silico functional predictions of fine-mapped OUD risk variants using the sequence-to-function deep-learning model AlphaGenome. (A) Deep-learning predictions for the *DDX6* locus regulatory variant (chr11:118713914:AT>A), illustrating the targeted downregulation of *FOXR1* expression specifically within the nucleus accumbens basal ganglia (log-fold change = - 0.166; quantile = -0.9999). (B) Predicted transcriptomic consequences of the *IL6R* variant (chr1:154411947:CT>C), demonstrating significant alterations in alternative splicing, including shifts in specific splice junctions (raw score = 1.127), overall splice site usage, and donor splice site probability within peripheral skeletal muscle. (C) Functional annotation of the protective *OPRM1* variant rs1799971 (chr6:154039662:A>G), showing a consistent modeled upregulation of *OPRM1* expression across critical reward and cognitive pathways, including the amygdala, nucleus accumbens, anterior cingulate cortex, dorsolateral prefrontal cortex, and hippocampus (log-fold change = 0.034; quantile = 0.9996).

In the Admixed American (AMR) cohort, the ASTN2 variant (chr9:116428405:C>T) was predicted to be associated with altered alternative splicing in the dorsolateral prefrontal cortex, affecting specific splice junctions (raw score = 0.933; quantile = 0.999) and donor splice site probability (raw score = 0.100; quantile = 0.989).

## Discussion

In this multi-ancestry genome-wide association study of the *All of Us* Research Program, we identified and replicated a novel risk locus at *DDX6*. The *DDX6* gene encodes a DEAD-box RNA helicase that is fundamentally involved in mRNA decapping, translational repression, and the assembly of processing bodies (P-bodies) and stress granules. Notably, our EUR GWAS identified a significant missense variant rs472419 (Ala314Pro/Thr) directly within this gene, suggesting a potential structural or functional alteration to the DDX6 protein itself. However, the genetic architecture at this locus appears complex; alongside this protein-coding variant, our *in silico* functional predictions utilizing AlphaGenome offer a localized hypothesis for a neighboring regulatory signal. Computational models suggest that the regulatory risk allele rs35994346 (chr11:118713914:AT>A) at this same locus may downregulate the *FOXR1* gene within the nucleus accumbens basal ganglia. *FOXR1* functions as a transcription factor critical for cellular stress resistance and neuronal survival, creating biological plausibility for OUD. If validated, its downregulation in the nucleus accumbens—a core node of the brain’s reward circuitry—would suggest a localized mechanism by which this locus might increase susceptibility to OUD, potentially by blunting cellular resilience to drug-induced or environmental stress. This would align well with the allostatic model of addiction which holds that mesolimbic dopaminergic function declines over time in response to repeated exposure to drugs, alcohol, or addictive behaviors such as gambling (49). Therefore, our findings might represent a previously undescribed genetic mediator of maladaptive neuroplasticity in addiction with plausible transdiagnostic importance. Future research should attempt to replicate the present findings among samples with other addiction disorders.

Furthermore, our findings successfully replicated previously established associations at *OPRM1* and *FURIN* in the EUR cohort, reinforcing their robust, cross-cohort roles in OUD etiology. Further, we replicated *OPRM1* in the AMR cohort and *FURIN* in the AFR cohort. The replication of the *OPRM1* variant rs1799971 (A118G) is particularly notable given its extensive history in both candidate-gene and genome-wide studies of addiction. While previous literature has debated the precise causal nature of this specific SNP, our deep-learning functional predictions offer a theoretical framework for its biological plausibility. AlphaGenome models predicted a consistent upregulation of *OPRM1* expression across a broad network of critical reward and cognitive pathways, including the amygdala, nucleus accumbens, anterior cingulate cortex, dorsolateral prefrontal cortex, and hippocampus. This modeled transcriptional enhancement of the primary target for opioids suggests a potential mechanism for how this allele may buffer against OUD liability.

Beyond *DDX6*, we identified several ancestry-specific loci of interest—including *IL6R*, and *SHISA9* in the EUR cohort; *GHR* in the AFR cohort; and *ASTN2* and *DCUN1D4* in the AMR cohort. Because these ancestry-specific signals did not replicate in independent datasets, the confidence with which we put them forward must be tempered. Nevertheless, prioritizing loci with established biological relevance offers compelling candidate regions for future exploratory research. For example, focusing on the EUR-associated *IL6R* locus, our GWAS identified a statistically robust signal comprising eight significant variants, anchored by the top variant rs7411976 (OR = 1.64). Notably, rs7411976 is in high linkage disequilibrium with the well-characterized functional missense variant rs2228145 (Asp358Ala). This variant alters the proteolytic cleavage site of the IL-6 receptor, enhancing its shedding from the cell surface. This structural shift elevates circulating soluble IL-6R while reducing membrane-bound receptors, effectively dampening classic pro-inflammatory signaling cascades. While it is compelling to consider how this established systemic immune modulation might influence addiction pathophysiology, our *in sili*co AlphaGenome model for a specific variant within this cluster (rs1179153554) pointed to alternative splicing alterations in peripheral skeletal muscle. This divergence highlights the highly pleiotropic nature of th*e IL*6R locus and underscores the need for tissue-specific experimental validation to determine whether this genetic risk for OUD is primarily mediated through its known immunological functions or via alternative, localized peripheral pathways.

Within the AMR cohort, while the *DCUN1D4* and *RNU6-1230P* variants exhibited substantial effect sizes (both with OR > 2.0), we prioritized *ASTN2* for further exploration due to its recognized role in glial-guided neuronal migration and the trafficking of synaptic surface proteins. Importantly, emerging literature provides strong support for its involvement in opioid-related phenotypes. Recent epigenetic studies of postmortem human brain tissue have demonstrated a significant convergence of altered histone acetylation near *ASTN2* specifically within the dorsolateral prefrontal cortex of opioid overdose cases, indicating that epigenetic dysregulation at this locus is tied to severe OUD outcomes (50). Furthermore, clinical genetic association studies have linked *ASTN2* single-nucleotide polymorphisms (e.g., rs7858836 and rs958804) to significant differences in postoperative fentanyl requirements, highlighting its potential role in modulating opioid sensitivity and analgesic response (51). Aligning closely with these independent epigenetic and clinical findings, our *in silico* data models a potential disruption of alternative splicing in *ASTN2* specifically within the dorsolateral prefrontal cortex for the AMR cohort.

A primary limitation of our study is the relatively small sample sizes for the AFR and AMR cohorts compared to the EUR group. This reduced statistical power likely explains the lack of replication for the novel AFR and AMR signals in the independent MVP cohort, which remains predominantly of European descent. Additionally, the reliance on EHR-derived phenotypes may introduce misclassification bias compared to structured clinical interviews. While it has been traditionally thought that the use of opioid-exposed controls reduces risk of confounding by opioid exposure opportunity, this assumption has recently been challenged (52). Finally, while our computational functional annotations provide a robust framework for mechanism discovery, these *in silico* predictions via AlphaGenome should not be interpreted as confirmed regulatory effects but as hypothesis-generating.

## Conclusion

This study expands our understanding of the genetic architecture of opioid use disorder by bridging the gap between multi-ancestry risk loci and their potential functional consequences. We identified and replicated a novel risk locus at *DDX6* alongside established signals at *OPRM1* and *FURIN*, providing compelling evidence for a role for all these genes in risk of OUD. Further, by utilizing advanced computational modeling, we were able to generate localized, biologically plausible hypotheses for their impacts on the brain’s reward circuitry. Furthermore, our *in silico* exploration of ancestry-specific variants underscores the critical need for continued investment in diverse genomic research. Integrating deep-learning functional annotations with diverse cohorts is essential to guide future experimental validation, ultimately ensuring that all populations benefit equitably from advances in precision medicine and targeted therapeutics for addiction.

## Data Availability

Data and Code Availability: The data supporting this study are available from the NIH All of Us Research Program. Analytic code is available from the authors upon reasonable request.

## Acknowledgments

The authors thank the participants and staff of the *All of Us* Research Program for their contributions. The *All of Us* Research Program is supported by the National Institutes of Health, Office of the Director: Regional Medical Centers: 1 OT2 OD026549; 1 OT2 OD026550; 1 OT2 OD026551; 1 OT2 OD026552; 1 OT2 OD026553; 1 OT2 OD026554; 1 OT2 OD026555; 1 OT2 OD026556; 1 OT2 OD026557; 1 OT2 OD026558; 1 OT2 OD026615; 1 OT2 OD025277; 1 OT2 OD025300; 1 OT2 OD025315; 1 OT2 OD025332; and 1 OT2 OD025276. In addition, the *All of Us* Research Program would not be possible without the partnership of its participants. This study was funded by an unrestricted grant from the Ohio Attorney General’s Office (#15974, Sprague).

## Data and Code Availability

The data supporting this study are available from the NIH *All of Us* Research Program. Analytic code is available from the authors upon reasonable request.

## Prior Dissemination

This article has been posted on the MedRxiv (DOI assignment pending).

## Footnotes

1 Note that the *All of Us* Research Program’s AMR designation includes individuals “who may be able to trace at least some of their distant ancestors back to North, Central, or South America” (39), and that 80.9% of our cases and 86.6% of our controls reported a Hispanic or Latino/a ethnicity (see Table 1 for details). AMR individuals are not necessarily members of Tribal Nations/communities, and they may have ancestral roots in Europe and/or Africa.

