## Supplement for "Genome-Wide Association Studies and Deep-Learning Functional Annotation of Opioid Use Disorder across Three Ancestries in the *All of Us* Research Program"

**Figure S1.** *Manhattan and quantile-quantile (Q-Q) plots for the European ancestry (EUR) analysis using All of Us (AoU)-provided principal components (PCs). λGC = genomic inflation factor.*

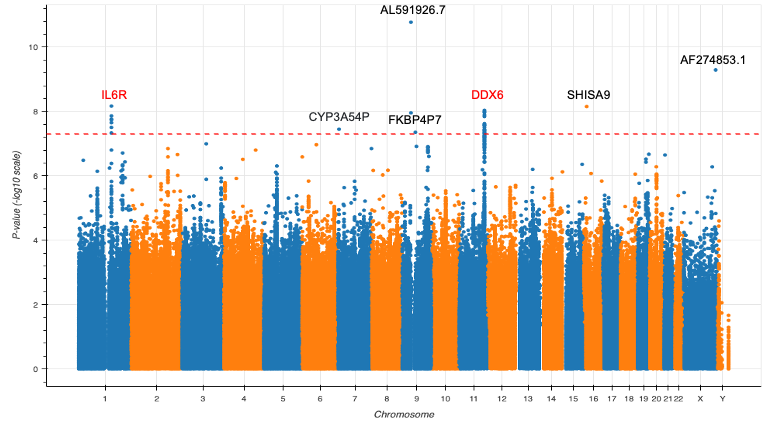

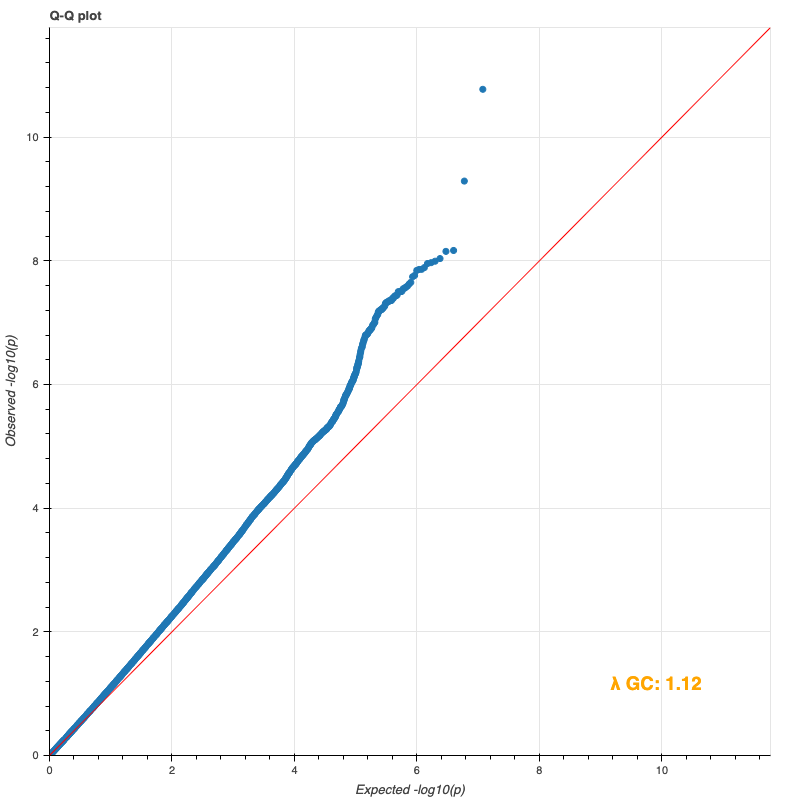

**Figure S2.** *Manhattan and quantile-quantile (Q-Q) plots for the African ancestry (AFR) analysis using All of Us (AoU)-provided principal components (PCs). λGC = genomic inflation factor.*

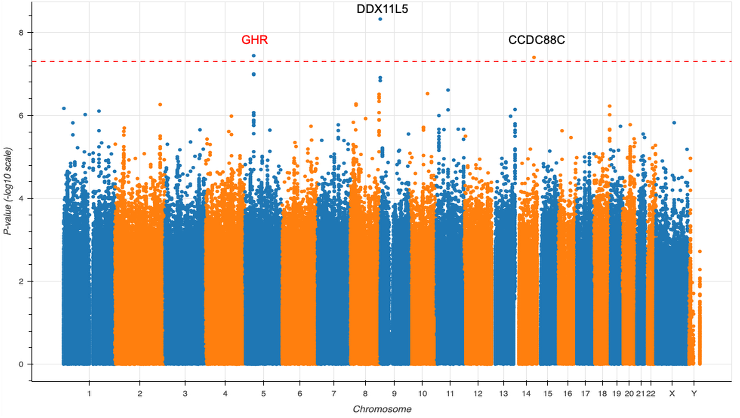

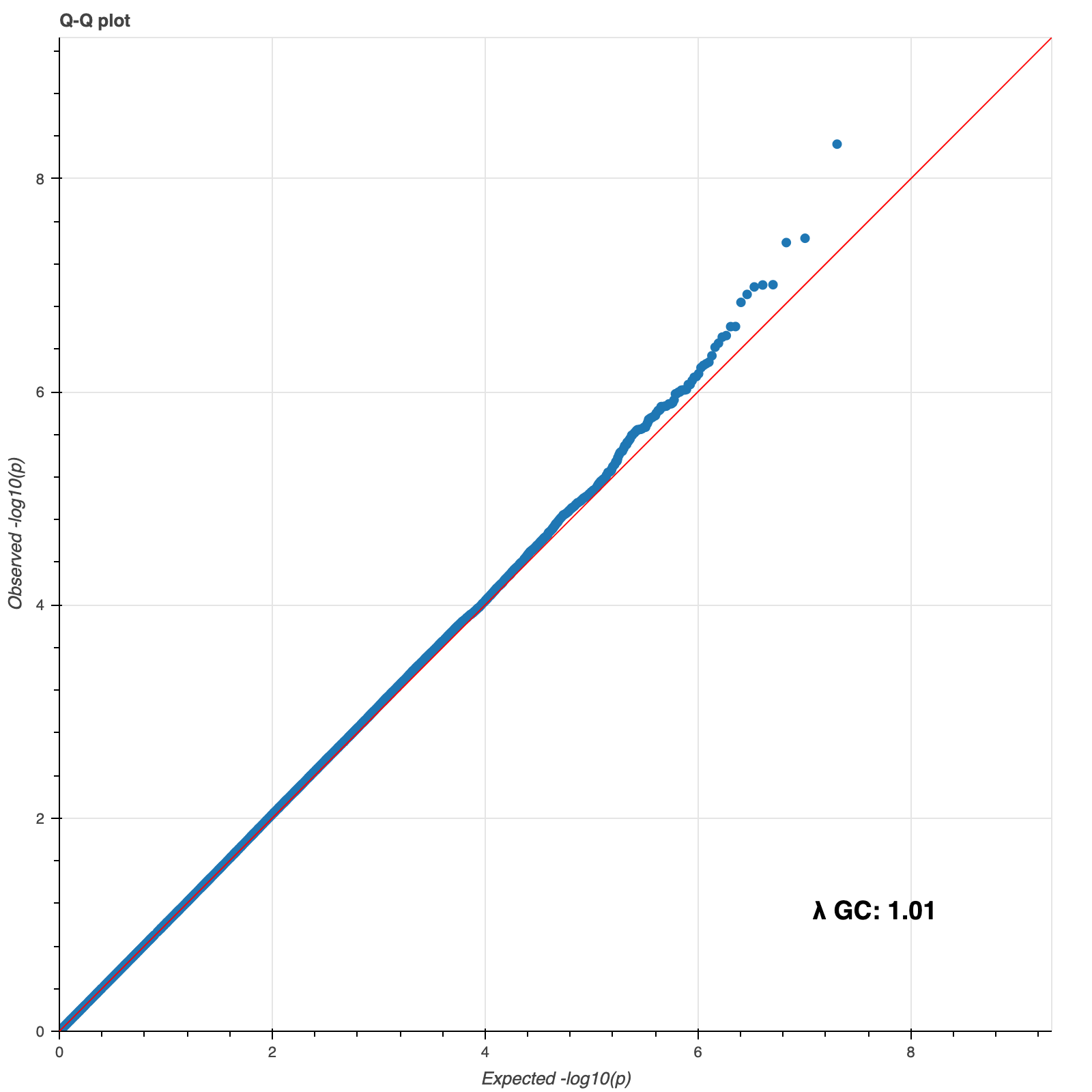

**Figure S3.** *Manhattan and quantile-quantile (Q-Q) plots for the Admixed American ancestry (AMR) analysis using ancestry-specific calculated principal components (PCs). λGC = genomic inflation factor.*

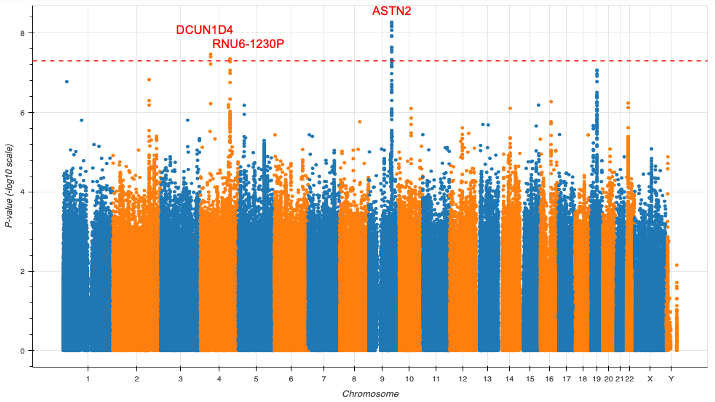

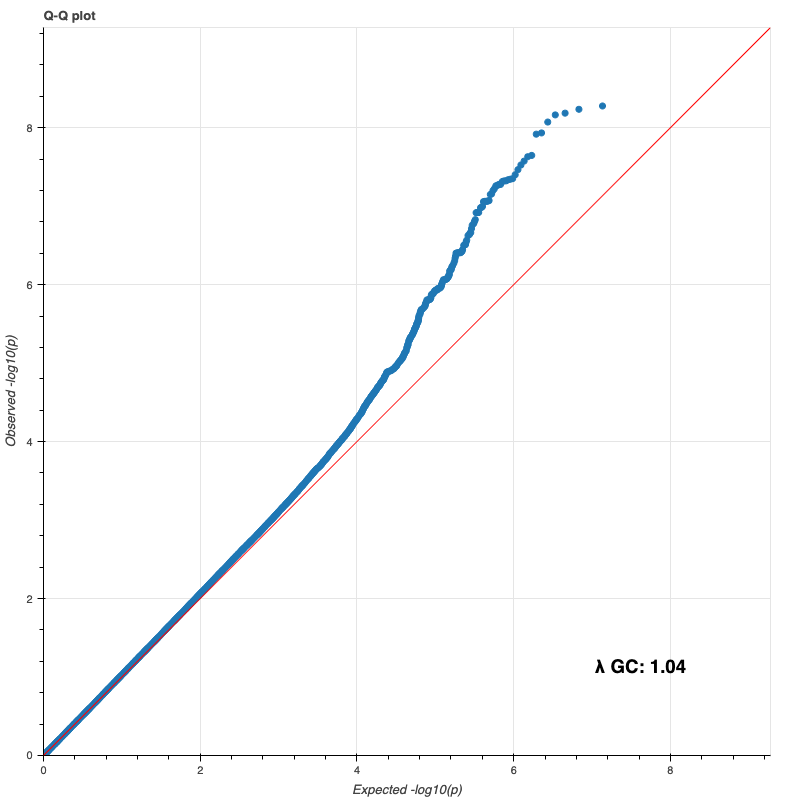

**Figure S4.** *Manhattan and quantile-quantile (Q-Q) plots for the European ancestry (EUR) analysis using ancestry-specific calculated principal components (PCs). λGC = genomic inflation factor.*

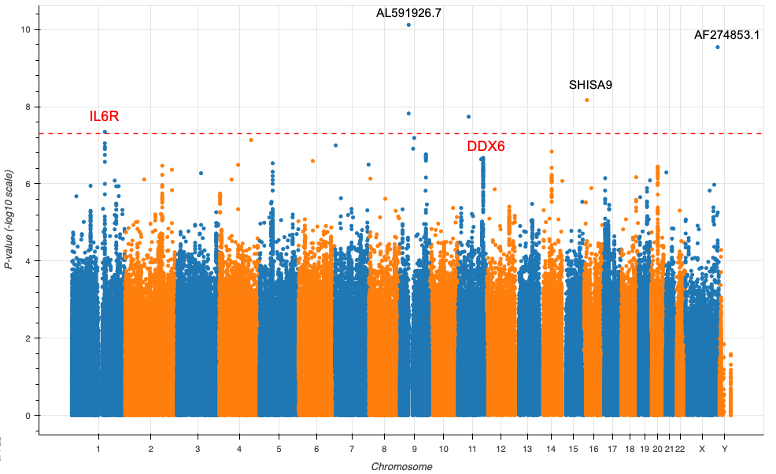

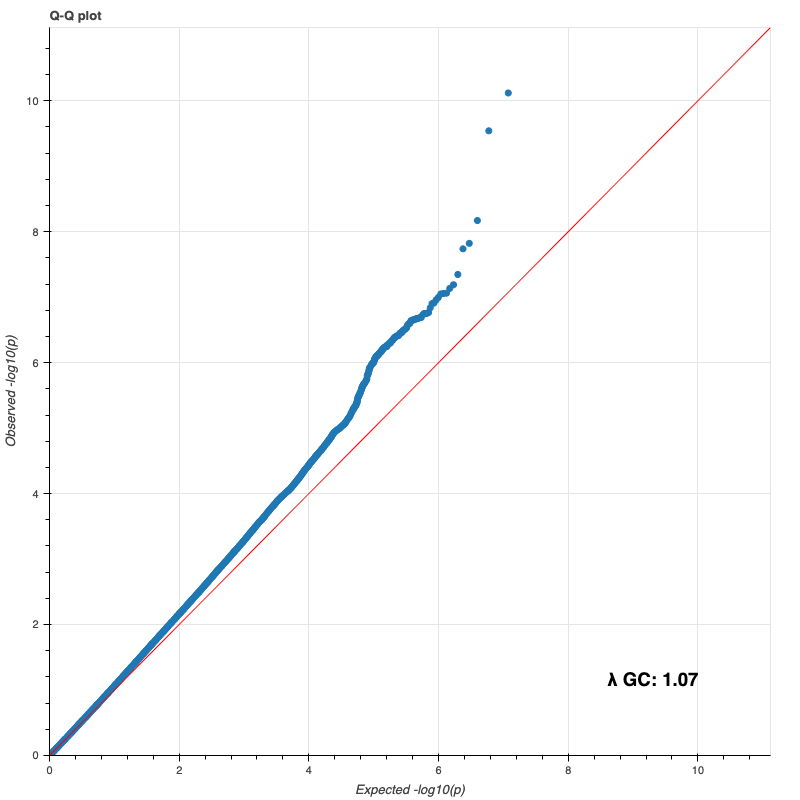

**Table S1.** *ICD-9/-10 codes used to identify opioid use disorder cases and exclude affected individuals from the control group.*

| **Domain** | **Code** | **Name** |
| --- | --- | --- |
| ICD-9 | 304.00 | Opioid type dependence, Unspecified |
|  | 304.01 | Opioid type dependence, Continuous |
|  | 304.02 | Opioid type dependence Episodic |
|  | 304.03 | Opioid type dependence in remission |
|  | 304.70 | Combinations of opioid type drug with any other drug dependence, unspecified |
|  | 304.71 | Combinations of opioid type drug with any other drug dependence, continuous |
|  | 304.72 | Combinations of opioid type drug with any other drug dependence, episodic use |
|  | 304.73 | Combinations of opioid type drug with any other drug dependence, in remission |
|  | 305.50 | Opioid abuse Unspecified |
|  | 305.51 | Opioid abuse Continuous |
|  | 305.52 | Opioid abuse Episodic |
|  | 305.53 | Opioid abuse in remission |
| ICD-10 | F11.10 | Opioid abuse uncomplicated |
|  | F11.11 | Opioid abuse in remission |
|  | F11.120 | Opioid abuse with intoxication uncomplicated |
|  | F11.121 | Opioid abuse with intoxication delirium |
|  | F11.122 | Opioid abuse with intoxication with perceptual disturbance |
|  | F11.129 | Opioid abuse with intoxication unspecified |
|  | F11.14 | Opioid abuse with opioid-induced mood disorder |
|  | F11.150 | Opioid abuse with opioid-induced psychotic disorder with delusions |
|  | F11.151 | Opioid abuse with opioid-induced psychotic disorder with hallucinations |
|  | F11.159 | Opioid abuse with opioid-induced psychotic disorder unspecified |
|  | F11.181 | Opioid abuse with opioid-induced sexual dysfunction |
|  | F11.182 | Opioid abuse with opioid-induced sleep disorder |
|  | F11.188 | Opioid abuse with other opioid-induced disorder |
|  | F11.19 | Opioid abuse with unspecified opioid-induced disorder |
|  | F11.20 | Opioid dependence uncomplicated |
|  | F11.21 | Opioid dependence in remission |
|  | F11.220 | Opioid dependence with intoxication uncomplicated |
|  | F11.221 | Opioid dependence with intoxication delirium |
|  | F11.222 | Opioid dependence with intoxication with perceptual disturbance |
|  | F11.229 | Opioid dependence with intoxication unspecified |
|  | F11.23 | Opioid dependence with withdrawal |
|  | F11.24 | Opioid dependence with opioid-induced mood disorder |
|  | F11.250 | Opioid dependence with opioid-induced psychotic disorder with delusions |
|  | F11.251 | Opioid dependence with opioid-induced psychotic disorder with hallucinations |
|  | F11.259 | Opioid dependence with opioid-induced psychotic disorder unspecified |
|  | F11.281 | Opioid dependence with opioid-induced sexual dysfunction |
|  | F11.282 | Opioid dependence with opioid-induced sleep disorder |
|  | F11.288 | Opioid dependence with other opioid-induced disorder |
|  | F11.29 | Opioid dependence with unspecified opioid-induced disorder |

*Note.* ICD-9 = International Classification of Diseases, Ninth Revision; ICD-10 = International Classification of Diseases, Tenth Revision

**Table S2.** *Descriptive statistics of lifetime substance use stratified by genetic ancestry and opioid use disorder among matched cases and controls.*

|  | **EUR**, *N* = 27,834 | | **AFR**, *N* = 15,103 | | **AMR**, *N* = 9,420 | |
| --- | --- | --- | --- | --- | --- | --- |
| **Lifetime Substance Use** | **Control (OUD -)**  *n* = 23,109 | **Case (OUD +)**  *n* = 4,725 | **Control (OUD -)**  *n* = 12,486 | **Case (OUD +)**  *n* = 2,617 | **Control (OUD -)**  *n* = 7,850 | **Case (OUD +)**  *n* = 1,570 |
| Cannabis | 13,757 (59.5%) | 3,465 (73.3%) | 6,426 (51.5%) | 1,614 (61.7%) | 3,349 (42.7%) | 1,044 (66.5%) |
| Cocaine | 4,529 (19.6%) | 2,598 (55.0%) | 2,594 (20.8%) | 1,185 (45.3%) | 1,431 (18.2%) | 754 (48.0%) |
| Methamphetamine | 2,205 (9.5%) | 1,733 (36.7%) | 355 (2.8%) | 179 (6.8%) | 689 (8.8%) | 407 (25.9%) |
| Prescription stimulants | 2,762 (12.0%) | 1,738 (36.8%) | 560 (4.5%) | 244 (9.3%) | 446 (5.7%) | 303 (19.3%) |
| Sedatives | 2,610 (11.3%) | 1,854 (39.2%) | 581 (4.7%) | 330 (12.6%) | 458 (5.8%) | 353 (22.5%) |
| Prescription opioids | 2,568 (11.1%) | 2,165 (45.8%) | 691 (5.5%) | 419 (16.0%) | 558 (7.1%) | 469 (29.9%) |
| Street opioids | 750 (3.2%) | 1,979 (41.9%) | 362 (2.9%) | 760 (29.0%) | 191 (2.4%) | 594 (37.8%) |
| Hallucinogens | 4,081 (17.7%) | 1,777 (37.6%) | 548 (4.4%) | 232 (8.9%) | 687 (8.8%) | 340 (21.7%) |
| Inhalants | 1,588 (6.9%) | 784 (16.6%) | 164 (1.3%) | 77 (2.9%) | 276 (3.5%) | 148 (9.4%) |
| None of these drugs | 7,336 (31.7%) | 591 (12.5%) | 4,264 (34.2%) | 373 (14.3%) | 3,610 (46.0%) | 226 (14.4%) |
| *Note.* Data displayed as counts and percentages. Percentages were calculated using the displayed *n* from the full sample used for GWAS as the denominator (after collapsing missing observations into non-endorsement to comply with the All of Us Research Program Data and Statistics Dissemination Policy, which does not permit dissemination of cells with raw counts < 20). While this approach is not ideal for presenting descriptive statistics, we confirmed that it had a negligible impact on percentages (due to the large sample sizes and because the motivation for doing this in the first place was because *n* < 20 for missingness on some variables). GWAS = genome-wide association study; OUD = opioid use disorder; EUR = European ancestry; AFR = African ancestry; AMR = Admixed American ancestry. “None of these drugs” was a separate response option. Past-three-month use was not presented due to documented errors in All of Us, making these variables unusable. | | | | | | |

**Table S3.** *Descriptive statistics of sociodemographic and health variables stratified by genetic ancestry and opioid use disorder among matched cases and controls.*

|  | **EUR**, *N* = 27,834 | | **AFR**, *N* = 15,103 | | **AMR**, *N* = 9,420 | |
| --- | --- | --- | --- | --- | --- | --- |
| **Sociodemographic and Health Variables** | **Control (OUD -)**  *n* = 23,109 | **Case (OUD +)**  *n* = 4,725 | **Control (OUD -)**  *n* = 12,486 | **Case (OUD +)**  *n* = 2,617 | **Control (OUD -)**  *n* = 7,850 | **Case (OUD +)**  *n* = 1,570 |
| **Education Level: Highest Grade Completed** |  |  |  |  |  |  |
| Highest Grade: 8th Grade or Less | 135 (0.6%) | 123 (2.7%) | 314 (2.6%) | 82 (3.4%) | 962 (12.6%) | 128 (8.5%) |
| Highest Grade: Nine Through Eleven | 524 (2.3%) | 462 (10.0%) | 1,490 (12.6%) | 526 (21.6%) | 962 (12.6%) | 344 (22.9%) |
| Highest Grade: Twelve Or GED | 3,245 (14.2%) | 1,589 (34.4%) | 3,991 (33.6%) | 997 (41.0%) | 2,192 (28.7%) | 579 (38.5%) |
| Highest Grade: College One to Three | 6,270 (27.4%) | 1,682 (36.4%) | 3,682 (31.0%) | 642 (26.4%) | 2,044 (26.8%) | 365 (24.3%) |
| Highest Grade: College Graduate | 6,518 (28.5%) | 487 (10.6%) | 1,501 (12.6%) | 149 (6.1%) | 963 (12.6%) | 67 (4.5%) |
| Highest Grade: Advanced Degree | 6,160 (27.0%) | 272 (5.9%) | 888 (7.5%) | 36 (1.5%) | 509 (6.7%) | 22 (1.5%) |
| **Marital Status: Current Marital Status** |  |  |  |  |  |  |
| Current Marital Status: Never Married | 4,134 (18.1%) | 1,600 (35.1%) | 4,587 (38.6%) | 1,082 (44.0%) | 2,022 (26.8%) | 605 (40.5%) |
| Current Marital Status: Living With Partner | 1,645 (7.2%) | 381 (8.4%) | 653 (5.5%) | 166 (6.7%) | 837 (11.1%) | 148 (9.9%) |
| Current Marital Status: Married | 12,710 (55.7%) | 940 (20.6%) | 2,821 (23.8%) | 344 (14.0%) | 2,990 (39.7%) | 250 (16.7%) |
| Current Marital Status: Separated | 454 (2.0%) | 275 (6.0%) | 821 (6.9%) | 239 (9.7%) | 427 (5.7%) | 147 (9.8%) |
| Current Marital Status: Divorced | 3,091 (13.5%) | 1,065 (23.3%) | 2,326 (19.6%) | 444 (18.0%) | 1,030 (13.7%) | 277 (18.6%) |
| Current Marital Status: Widowed | 791 (3.5%) | 301 (6.6%) | 662 (5.6%) | 186 (7.6%) | 234 (3.1%) | 66 (4.4%) |
| **Active Duty: Lifetime Active Duty Service** |  |  |  |  |  |  |
| Lifetime Active Duty Service: No | 20,744 (90.5%) | 4,156 (90.1%) | 10,858 (88.9%) | 2,238 (89.0%) | 7,233 (93.7%) | 1,421 (93.1%) |
| Lifetime Active Duty Service: Yes | 2,169 (9.5%) | 459 (9.9%) | 1,360 (11.1%) | 277 (11.0%) | 487 (6.3%) | 106 (6.9%) |
| **Insurance: Current Health Insurance** |  |  |  |  |  |  |
| Current Health Insurance: No | 642 (2.8%) | 373 (8.2%) | 1,230 (10.3%) | 177 (7.1%) | 732 (9.7%) | 78 (5.1%) |
| Current Health Insurance: Yes | 22,170 (97.2%) | 4,193 (91.8%) | 10,748 (89.7%) | 2,325 (92.9%) | 6,796 (90.3%) | 1,439 (94.9%) |
| **Employment: Current Employment Status** |  |  |  |  |  |  |
| Employment Status: Employed For Wages | 12,226 (52.9%) | 604 (12.8%) | 3,589 (28.7%) | 216 (8.3%) | 3,383 (43.1%) | 191 (12.2%) |
| Employment Status: Self Employed | 1,703 (7.4%) | 232 (4.9%) | 695 (5.6%) | 108 (4.1%) | 566 (7.2%) | 92 (5.9%) |
| Employment Status: Student/Other Response | 969 (4.2%) | 288 (6.1%) | 819 (6.6%) | 190 (7.3%) | 550 (7.0%) | 122 (7.8%) |
| Employment Status: Homemaker | 815 (3.5%) | 134 (2.8%) | 257 (2.1%) | 79 (3.0%) | 477 (6.1%) | 62 (3.9%) |
| Employment Status: Retired | 3,474 (15.0%) | 614 (13.0%) | 1,955 (15.7%) | 336 (12.8%) | 553 (7.0%) | 92 (5.9%) |
| Employment Status: Out Of Work (< One Year) | 761 (3.3%) | 518 (11.0%) | 737 (5.9%) | 208 (7.9%) | 498 (6.3%) | 174 (11.1%) |
| Employment Status: Out Of Work (≥ One Year) | 863 (3.7%) | 709 (15.0%) | 1,075 (8.6%) | 407 (15.6%) | 667 (8.5%) | 298 (19.0%) |
| Employment Status: Unable To Work | 2,298 (9.9%) | 1,626 (34.4%) | 3,359 (26.9%) | 1,073 (41.0%) | 1,156 (14.7%) | 539 (34.3%) |
| **Income: Current Annual Household Income** |  |  |  |  |  |  |
| Annual Income: less than $10k | 1,647 (8.0%) | 1,471 (38.4%) | 3,450 (38.1%) | 1,145 (60.1%) | 1,136 (22.9%) | 563 (53.3%) |
| Annual Income: $10k-$25k | 2,060 (10.1%) | 1,010 (26.4%) | 2,044 (22.5%) | 470 (24.7%) | 1,040 (21.0%) | 249 (23.6%) |
| Annual Income: $25k-$35k | 1,458 (7.1%) | 382 (10.0%) | 934 (10.3%) | 135 (7.1%) | 658 (13.3%) | 88 (8.3%) |
| Annual Income: $35k-$50k | 1,892 (9.2%) | 301 (7.9%) | 830 (9.2%) | 88 (4.6%) | 573 (11.6%) | 64 (6.1%) |
| Annual Income: $50k-$75k | 2,958 (14.5%) | 278 (7.3%) | 782 (8.6%) | 38 (2.0%) | 595 (12.0%) | 49 (4.6%) |
| Annual Income: $75k or More | 10,454 (51.1%) | 384 (10.0%) | 1,027 (11.3%) | 28 (1.5%) | 949 (19.2%) | 44 (4.2%) |
| **Home Own: Current Home Own** |  |  |  |  |  |  |
| Current Home Own: Own | 13,899 (61.7%) | 949 (21.8%) | 2,662 (23.0%) | 162 (6.8%) | 2,033 (27.7%) | 169 (12.0%) |
| Current Home Own: Rent | 6,577 (29.2%) | 2,031 (46.7%) | 7,282 (62.8%) | 1,708 (71.5%) | 4,402 (59.9%) | 800 (56.7%) |
| Current Home Own: Other Arrangement | 2,041 (9.1%) | 1,368 (31.5%) | 1,652 (14.2%) | 519 (21.7%) | 908 (12.4%) | 442 (31.3%) |
| **Living Situation: Past-Six-Month Housing** |  |  |  |  |  |  |
| Stable House Concern: No | 19,749 (86.4%) | 2,365 (51.2%) | 8,959 (73.2%) | 1,547 (60.8%) | 5,914 (77.7%) | 788 (51.7%) |
| Stable House Concern: Yes | 3,120 (13.6%) | 2,257 (48.8%) | 3,284 (26.8%) | 999 (39.2%) | 1,697 (22.3%) | 736 (48.3%) |
| **Health Advice: Spoken To/Seen a Mental Health Professional (During Past 12 Months)** |  |  |  |  |  |  |
| Spoken To Mental Health Professional: No | 9,159 (67.5%) | 468 (36.4%) | 2,223 (72.4%) | 172 (49.9%) | 1,702 (73.3%) | 116 (43.4%) |
| Spoken To Mental Health Professional: Yes | 4,406 (32.5%) | 816 (63.6%) | 846 (27.6%) | 173 (50.1%) | 619 (26.7%) | 151 (56.6%) |
| **Overall Health: General Mental Health** |  |  |  |  |  |  |
| General Mental Health: Poor | 782 (3.4%) | 478 (10.3%) | 470 (3.9%) | 187 (7.4%) | 299 (3.9%) | 164 (10.7%) |
| General Mental Health: Fair | 3,420 (15.0%) | 1,397 (30.2%) | 2,378 (19.6%) | 720 (28.6%) | 1,253 (16.2%) | 463 (30.3%) |
| General Mental Health: Good | 6,715 (29.4%) | 1,488 (32.2%) | 3,792 (31.2%) | 847 (33.6%) | 2,651 (34.3%) | 460 (30.1%) |
| General Mental Health: Very Good | 7,846 (34.3%) | 851 (18.4%) | 3,023 (24.9%) | 448 (17.8%) | 2,062 (26.7%) | 279 (18.3%) |
| General Mental Health: Excellent | 4,113 (18.0%) | 405 (8.8%) | 2,494 (20.5%) | 316 (12.5%) | 1,472 (19.0%) | 161 (10.5%) |
| **Overall Health: General Quality of Life** |  |  |  |  |  |  |
| General Quality: Poor | 605 (2.7%) | 544 (11.8%) | 439 (3.6%) | 155 (6.2%) | 245 (3.2%) | 167 (11.0%) |
| General Quality: Fair | 2,532 (11.1%) | 1,334 (29.1%) | 2,512 (20.8%) | 778 (31.4%) | 1,465 (19.0%) | 488 (32.3%) |
| General Quality: Good | 6,517 (28.6%) | 1,685 (36.7%) | 4,656 (38.6%) | 951 (38.3%) | 3,146 (40.8%) | 520 (34.4%) |
| General Quality: Very Good | 8,910 (39.0%) | 742 (16.2%) | 3,015 (25.0%) | 378 (15.2%) | 1,887 (24.5%) | 224 (14.8%) |
| General Quality: Excellent | 4,257 (18.7%) | 287 (6.3%) | 1,433 (11.9%) | 219 (8.8%) | 965 (12.5%) | 114 (7.5%) |
| **Overall Health: General Physical Health** |  |  |  |  |  |  |
| General Physical Health: Poor | 1,331 (5.8%) | 754 (16.4%) | 858 (7.1%) | 286 (11.5%) | 680 (8.8%) | 239 (15.7%) |
| General Physical Health: Fair | 4,574 (20.0%) | 1,599 (34.8%) | 3,696 (30.5%) | 915 (36.7%) | 2,171 (28.1%) | 580 (38.2%) |
| General Physical Health: Good | 8,159 (35.7%) | 1,492 (32.4%) | 4,533 (37.5%) | 843 (33.9%) | 2,892 (37.5%) | 447 (29.4%) |
| General Physical Health: Very Good | 6,797 (29.7%) | 612 (13.3%) | 2,162 (17.9%) | 292 (11.7%) | 1,434 (18.6%) | 188 (12.4%) |
| General Physical Health: Excellent | 1,999 (8.7%) | 143 (3.1%) | 853 (7.0%) | 154 (6.2%) | 544 (7.0%) | 66 (4.3%) |
| **Overall Health: Average Fatigue (Past Week)** |  |  |  |  |  |  |
| Average Fatigue 7 Days: None | 3,576 (15.6%) | 360 (7.8%) | 2,649 (21.7%) | 450 (17.7%) | 1,583 (20.5%) | 206 (13.4%) |
| Average Fatigue 7 Days: Mild | 9,726 (42.5%) | 1,199 (25.9%) | 4,091 (33.5%) | 749 (29.5%) | 2,377 (30.8%) | 413 (26.8%) |
| Average Fatigue 7 Days: Moderate | 7,007 (30.6%) | 1,878 (40.5%) | 4,056 (33.2%) | 960 (37.9%) | 2,577 (33.4%) | 538 (34.9%) |
| Average Fatigue 7 Days: Severe | 2,118 (9.2%) | 959 (20.7%) | 1,104 (9.0%) | 295 (11.6%) | 932 (12.1%) | 289 (18.8%) |
| Average Fatigue 7 Days: Very Severe | 479 (2.1%) | 242 (5.2%) | 304 (2.5%) | 82 (3.2%) | 256 (3.3%) | 95 (6.2%) |
| **Overall Health: Emotional Prob. (Past Week)** |  |  |  |  |  |  |
| Emotional Problem 7 Days: Never | 4,072 (17.8%) | 419 (9.1%) | 3,154 (26.0%) | 430 (17.0%) | 2,013 (26.2%) | 187 (12.2%) |
| Emotional Problem 7 Days: Rarely | 7,010 (30.6%) | 738 (16.0%) | 2,802 (23.1%) | 463 (18.3%) | 1,830 (23.8%) | 236 (15.5%) |
| Emotional Problem 7 Days: Sometimes | 7,499 (32.8%) | 1,512 (32.7%) | 4,061 (33.4%) | 989 (39.2%) | 2,407 (31.3%) | 535 (35.0%) |
| Emotional Problem 7 Days: Often | 3,280 (14.3%) | 1,328 (28.7%) | 1,556 (12.8%) | 466 (18.4%) | 1,049 (13.6%) | 382 (25.0%) |
| Emotional Problem 7 Days: Always | 1,013 (4.4%) | 623 (13.5%) | 581 (4.8%) | 178 (7.0%) | 388 (5.0%) | 187 (12.2%) |
| **Disability: Difficulty Concentrating, Remembering, or Making Decisions** |  |  |  |  |  |  |
| Difficulty Concentrating, etc.: No | 14,103 (86.7%) | 1,362 (64.5%) | 4,711 (83.3%) | 593 (69.7%) | 3,743 (85.6%) | 386 (67.5%) |
| Difficulty Concentrating, etc.: Yes | 2,165 (13.3%) | 751 (35.5%) | 945 (16.7%) | 258 (30.3%) | 628 (14.4%) | 186 (32.5%) |
| **Treated with Respect by Healthcare Provider** |  |  |  |  |  |  |
| Respected By Provider: Always | 10,448 (70.9%) | 822 (56.3%) | 2,626 (71.7%) | 282 (68.6%) | 1,888 (72.6%) | 195 (63.9%) |
| Respected By Provider: Most Of The Time | 3,601 (24.4%) | 454 (31.1%) | 720 (19.7%) | 73 (17.8%) | 518 (19.9%) | 67 (22.0%) |
| Respected By Provider: Some of The Time, None of the Time, or Other Response | 689 (4.7%) | 184 (12.6%) | 315 (8.6%) | 56 (13.6%) | 194 (7.5%) | 43 (14.1%) |
| **Average pain (0-10) (Past Week)** |  |  |  |  |  |  |
| Mean (SD) | 3.16 (2.77) | 5.58 (2.70) | 4.8 (3.2) | 5.7 (3.1) | 4.7 (3.3) | 6.3 (2.9) |
| Median [Q1, Q3] | 2.00 [1.00, 5.00] | 6.00 [4.00, 8.00] | 5.0 [2.0, 7.0] | 6.0 [4.0, 8.0] | 5.0 [2.0, 7.0] | 7.0 [5.0, 8.0] |
| *Note.* Data displayed as counts and percentages, unless otherwise indicated. Some cells were collapsed to comply with the All of Us Research Program Data and Statistics Dissemination Policy, which does not permit dissemination of cells with raw counts < 20. When noted, “Other Response” comprises “Don’t Know”, “Skip”, “Prefer Not to Answer”, and related responses; otherwise, these were set to missing, and counts/percentages for missing data were suppressed. Some percentages do not sum to 100% due to rounding. Percentages were calculated using observed (non-missing) survey *n* as the denominator, whereas the *n* from the full sample used for the GWAS is displayed for each level of stratification as reference. SD = standard deviation; Q1 = first quartile; Q3 = third quartile; OUD = opioid use disorder; EUR = European ancestry; AFR = African ancestry; AMR = Admixed American ancestry. For interested readers who would like to directly reference the All of Us questions and response options, the concept codes are: 1585940, 1585852, 1585892, 1585386, 903575, 1585952, 1585375, 1585370, 1585886, 1585717, 1585723, 1585729, 1585747, 1585748, 1585760, 43530402, 43530439 (<https://databrowser.researchallofus.org/>). | | | | | | |
